# Functional Evidence for Substantia Nigra Pars Reticulata Involvement in Auditory Verbal Hallucinations in Treatment-Resistant Schizophrenia

**DOI:** 10.1101/2025.05.12.25327399

**Authors:** Anruo Shen, Yousef Salimpour, Ankur Butala, Min Jae Kim, Ki Sueng Choi, Michael Bray, Frederick Nucifora, David Schretlen, Philip Harvey, William S. Anderson, Martijn Figee, Kelly A. Mills, Akira Sawa, Nicola G. Cascella

## Abstract

Auditory verbal hallucinations (AVH) in treatment-resistant schizophrenia (TR SZ) are often unresponsive to pharmacologic interventions, necessitating novel therapeutic approaches. We report a case of a patient with persistent AVH who underwent deep brain stimulation (DBS) of the substantia nigra pars reticulata (SNr). To investigate the neurophysiological effects of SNr stimulation on cortical activity during AVH, we performed intraoperative electrocorticography (ECoG) over the left inferior parietal cortex using a 63-channel grid. Real-time recordings captured episodes of AVH and revealed elevated theta-gamma phase-amplitude coupling (PAC), a marker of aberrant cortical synchronization. DBS intervention resulted in normalization of PAC dynamics, with both average PAC and spatial distribution returning to non-AVH levels. Clinically, SNr DBS was associated with a 64% reduction in AVH frequency. These findings provide the first real-time functional evidence linking SNr activity to cortical oscillatory changes underlying AVH. This case supports the potential of SNr as a novel neuromodulatory target in TR SZ and highlights theta-gamma PAC as a candidate biomarker for mechanistic tracking of AVH symptomatology.

---

We have previously reported that deep brain stimulation (DBS) targeting the substantia nigra pars reticulata (SNr) in a patient with treatment-resistant schizophrenia (TR SZ) provided immediate and sustained relief from auditory verbal hallucinations (AVH) that was unresponsive to pharmacologic treatment (1). To investigate the effect of SNr stimulation on pathological neurophysiologic phenomena associated with AVH in TR SZ, we analyzed, in a new patient, theta-gamma phase-amplitude coupling (PAC), a key feature of hierarchical processing and dynamic communication between neural circuits, particularly in cognition and perception.

AVHs are thought to involve dysfunction within auditory and language networks (2) and theta-gamma PAC has been associated with auditory function. In schizophrenia, theta-gamma PAC shows less lateralization over auditory cortex relative to healthy controls (3). However, most PAC studies are based on Electroencephalography (EEG) (4–6), which has limited spatial resolution and signal fidelity, contributing to inconsistent findings. In contrast, our study employed Electrocorticography (ECoG) signals, which provide direct cortical recordings with higher spatial and temporal resolution. By focusing on the left inferior parietal cortex, including the postcentral, inferior parietal, angular and supramarginal gyri, as defined by the Automated Anatomical Labeling (AAL) atlas (7), we aimed to capture neural dynamics directly related to language processing, which is closely linked to AVH (8–13). Previous EEG studies have shown that the presence of AVH is associated with elevated theta-gamma PAC (14–16). Consistent with this, we observed elevated theta-gamma PAC during AVH episodes. Here we have further shown that when DBS was applied during AVH episodes, PAC dynamics were normalized, restoring both the average value of PAC and the distribution of synchronization to levels comparable to the non-AVH, non-DBS condition. This suggests a potential modulatory effect of DBS in restoring oscillatory coupling balance while treating AVH. This is the first reported case providing real-time functional evidence linking SNr activity to cortical oscillatory changes in AVH. SNr DBS acutely improves AVH which is associated with a down-regulation of theta-gamma PAC.

This study was registered on ClinicalTrials.gov (NCT02361554). The subject is a man in his 30s with TR SZ. His persistent AVH emerged when he was an adolescent. Antipsychotic medications, including clozapine, failed to significantly reduce them. Electroconvulsive therapy was never used. Prior to enrollment, the subject’s capacity to give written informed consent was independently assessed by a psychiatrist not involved in the study.

The implant of the electrodes was done under light anesthesia which was withheld prior to the ECoG recording. We used a temporary implanted 63-channel grid electrode over the left inferior parietal cortex involved in language hierarchical processing network during bilateral DBS lead placement procedure of the SNr (**Fig. 1A**). The brain electrodes were registered to the AAL atlas, as shown in **Fig. 1B**. Time-series ECoG data was collected intraoperatively for 11.5 min during which DBS was first turned-off for 6.5 min and then turned-on for 5 min, with patient providing real-time reports of AVH occurrence using a button press held during hallucinations and released when they abated. The influence of anesthesia is a potential limiting factor as illustrated in **Fig. 1C**, as some AVH episodes were reported with durations shorter than one second. Standard preprocessing steps were applied, including DBS artifact removal using independent component analysis (ICA) and DBSFILT toolbox (17), notch filtering at 60Hz to remove line noise, bandpass filtering (1–200 Hz), demeaning, and average re-referencing. Theta-gamma phase-amplitude coupling (PAC) were computed and compared using a control-variable approach.

**Figure 1.**
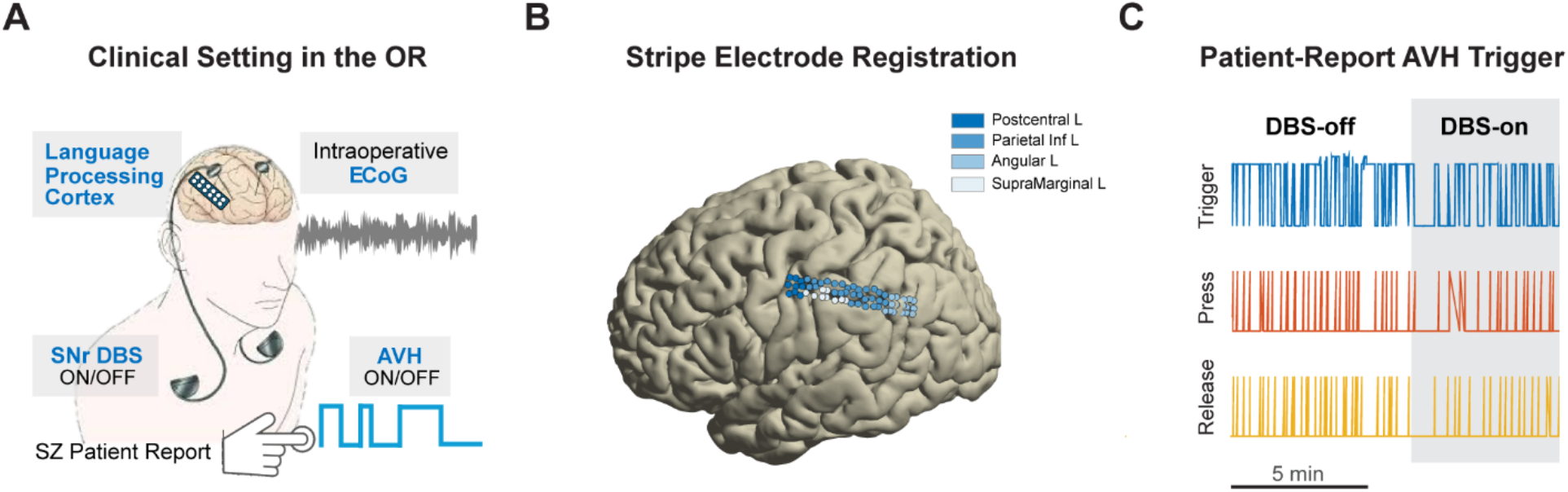
Experimental setup and patient-reported AVH dynamics during intraoperative DBS. (A) Intraoperative setup showing simultaneous SNr DBS off/on, language cortex ECoG recording, and real-time AVH reporting. (B) Electrode registration on left hemisphere cortical regions over the language processing areas: postcentral, parietal inferior, angular, and supramarginal gyri. (C) Patient-reported AVH events during DBS-off and DBS-on phases, indicating time-locked changes in AVH trigger, start, and end dynamics. AVH events were 64.34% more frequently reported during DBS-off periods.

SNr DBS was associated with a 64% reduction in self-reported AVH. Although the observed theta–gamma PAC magnitude was modest, a permutation test against 100 phase-amplitude mismatch surrogates confirmed that the coupling was statistically significant and non-random (p = 0.009; **Fig. 2A**). In addition, we identified three spatially localized regions with elevated PAC activity during AVH, situated in the inferior parietal lobe and postcentral gyrus (**Fig. 2B**), which were selected for further analysis. AVH appears to disrupt the balance of theta–gamma PAC, leading to an increased PAC across different regions **(Fig. 2C**). Regardless of DBS condition or location, mean PAC value during AVH increased, as shown in **Fig. 2D**, suggesting a synchronization at the language processing cortex during AVH. However, relative SNr DBS revealed a decreased baseline and theta-gamma PAC hypersynchronization.

**Figure 2.**
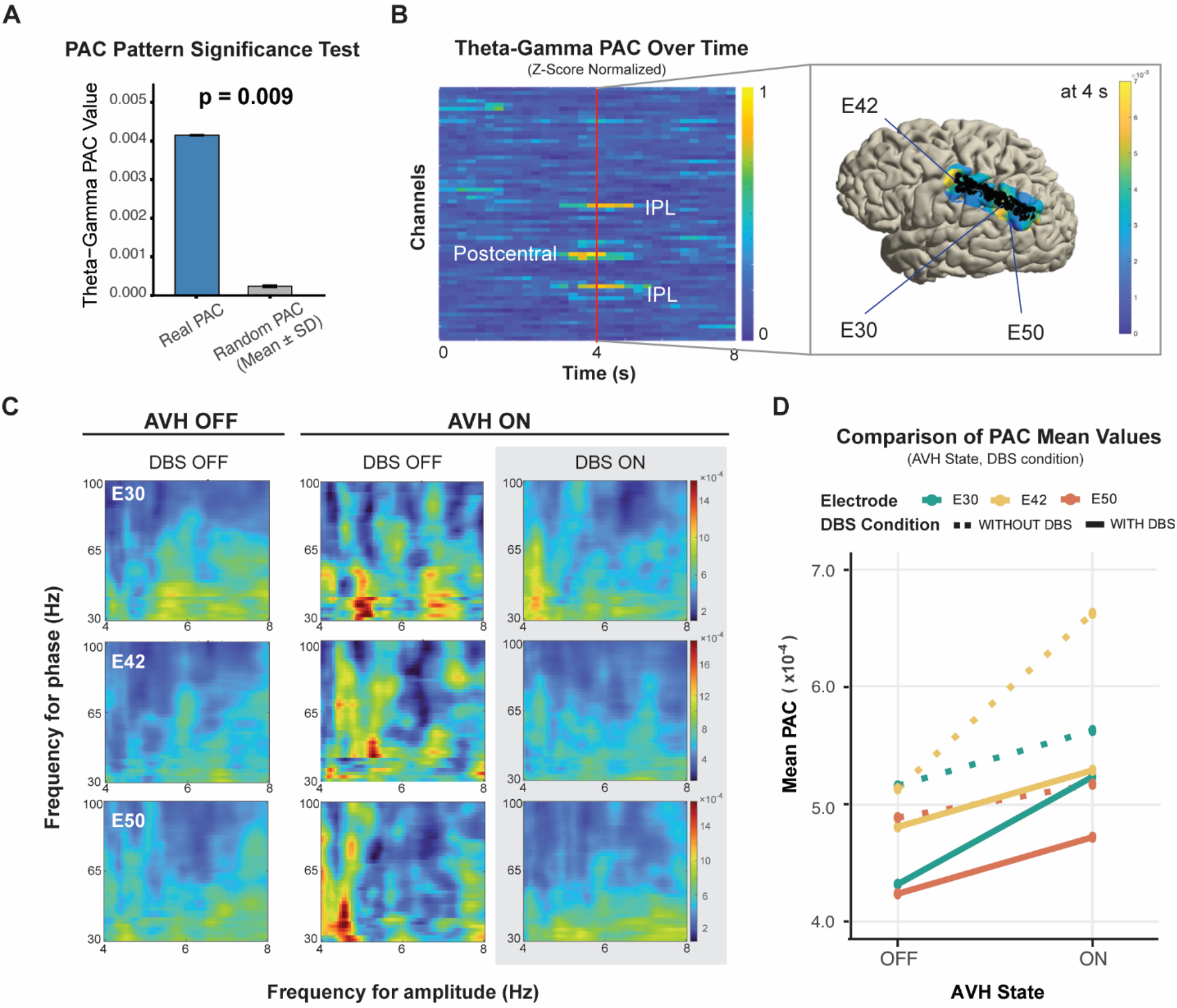
Theta-Gamma PAC dynamics across AVH states and DBS conditions. (A) Permutation test of maximal theta–gamma PAC values against 100 phase–amplitude mismatched surrogates revealed significant cross-frequency coupling (p = 0.009). (B) Z-scored temporal PAC map during an AVH episode, highlighting three hyperactive channels selected for analysis, located in inferior parietal lobe (IPL) and postcentral gyrus. Their locations were further illustrated in the brain mapping. (C) Theta-gamma PAC levels at electrodes E30, E42, and E50 reveal condition-dependent modulation of PAC. AVH appears to disrupt the balance of theta–gamma PAC, leading to an increased PAC across different regions. (D) Mean PAC increased during AVH episodes but was decreased by SNr DBS to levels comparable to the non-AVH, non-DBS condition.

While the precise mechanism of the impact of DBS on neurocircuits remains elusive (18), SNr DBS has shown preliminary promise in alleviating TR-AVH. GABAergic neurons of the SNr provide the primary output of the basal ganglia. SNr neurons fire spontaneously at 30–60 Hz in murine models, and the prevailing view is that they tonically inhibit thalamic neurons that return their excitatory output to the cortical regions of the brain (19). There is post-mortem evidence of GABAergic abnormalities in the SNr of SZ (20) including TR-SZ (21). It is conceivable that our stimulation overrides pathological firing patterns in the SNr resulting in a modulation of the increased theta-gamma PAC associated with AVH.

## Data Availability

All data produced in the present study are available upon reasonable request to the authors.

## Acknowledgements

We extend our gratitude to the participant and his family. We also thank Ms. Yukiko Lema for her suggestions for formatting the figures and her role in research management.

Dr. Sawa reports having received research funding from the National Institutes of Health Grant (No. P50MH136297). Dr. Cascella reports having received research funding from the National Institutes of Health Grant (No. U01MH130625).

